# A Phase 2/3 study of S-217622 in participants with SARS-CoV-2 infection (Phase 3 part)

**DOI:** 10.1101/2022.07.15.22277670

**Authors:** Hiroshi Yotsuyanagi, Norio Ohmagari, Yohei Doi, Takumi Imamura, Takuhiro Sonoyama, Genki Ichihashi, Takao Sanaki, Yuko Tsuge, Takeki Uehara, Hiroshi Mukae

## Abstract

**Background:** Limited treatment options exist for patients with mild-to-moderate coronavirus disease 2019 (COVID-19), irrespective of vaccination history or risk status. Ensitrelvir is a novel oral severe acute respiratory syndrome coronavirus 2 (SARS-CoV-2) 3C-like protease inhibitor. While phase 2 studies of ensitrelvir have demonstrated promising results in treating mild-to-moderate COVID-19, evaluation of its clinical efficacy due to shifting vaccination status and emergence of the Omicron variant represents significant challenges. Here, we describe the protocol for a phase 3 study designed to evaluate the efficacy and safety of ensitrelvir in patients with mild-to-moderate COVID-19, regardless of risk status or vaccination history.

**Methods:** This is a multicenter, randomized, double-blind, placebo-controlled, phase 3 study. Patients with mild-to-moderate COVID-19 within 120 hours from onset will be randomized in a 1:1:1 ratio into 3 treatment arms–ensitrelvir 125 mg (375 mg loading dose on Day 1), ensitrelvir 250 mg (750 mg loading dose on Day 1), and placebo. The study interventions will be administered orally, once daily, for 5 days. The primary endpoint will be the time to resolution of 5 symptoms of COVID-19 (stuffy or runny nose, sore throat, cough, feeling hot or feverish, and low energy or tiredness), and the key secondary endpoints will include the change from baseline on Day 4 in the amount of SARS-CoV-2 viral RNA and the time to first negative SARS-CoV-2 viral titer. The primary population for the primary and key secondary endpoints will be patients with <72 hours from COVID-19 onset to randomization and, subsequently, patients in entire patient population (<120 hours) in the ensitrelvir 125 mg group. Closed testing procedure will be used for the primary and key secondary endpoints in both the primary and entire patient populations. All safety assessments and adverse events will be reported.

**Discussion:** In a post hoc analysis of the phase 2b study, compared with placebo, ensitrelvir demonstrated a reduced time to resolution of 5 symptoms in patients with mild-to-moderate COVID-19. Through this study, we intend to validate and establish the efficacy and safety of ensitrelvir in patients with mild-to-moderate COVID-19.

**Trial registration:** Japan Registry of Clinical Trials (https://jrct.niph.go.jp): jRCT2031210350.

## 1. Introduction

Coronavirus disease 2019 (COVID-19) is an infectious respiratory illness caused by severe acute respiratory syndrome coronavirus 2 (SARS-CoV-2)^1^ and was declared a global pandemic by the World Health Organization (WHO) in March 2020.^2^ Globally, as of October 2022, 630 million confirmed cases of COVID-19 have been reported, with 6.6 million associated deaths.^3^ Common symptoms of COVID-19 include fever, cough, fatigue, shortness of breath, headache, body aches, sore throat, runny or stuffy nose, diarrhea, nausea, and vomiting; some patients may also experience loss of smell and/or taste.^4^ High-risk patients such as the elderly and those with comorbidities and underlying health conditions such as cardiovascular disease, respiratory disease, renal disease, diabetes mellitus, obesity, or immunodeficiency are at a greater risk of developing severe COVID-19 symptoms.^5, 6^ Some COVID-19 symptoms may persist even after cessation of SARS-CoV-2 shedding and recovery from acute illness, causing a wide range of health problems collectively known as the post-acute COVID-19 syndrome.^7, 8^

The SARS-CoV-2 Omicron is a variant of concern (VOC) designated by WHO.^9^ This variant is characterized by spike protein mutations that lead to increased transmissibility and is known to escape from SARS-CoV-2 neutralizing antibodies.^10^ A study from the United Kingdom indicated that common symptoms reported in patients infected with the Omicron variant of SARS-CoV-2 were mostly mild (runny nose, headache, sore throat, sneezing, persistent cough, and hoarse voice).^11^ The risk of hospitalization and hospitalization rates were lower during the prevalence of the Omicron variant compared with that of the Delta variant,^11, 12^ consistent with studies reporting that infection with the Omicron variant is less severe than that with the Delta variant.^13-15^ Although milder, several reports have indicated that infection with the Omicron variant is associated with a higher excess mortality than that with the Delta variant^16^ and seasonal influenza.^12^ In Japan, the first patient infected with the Omicron variant of SARS-CoV-2 was reported in November 2021. Subsequently, various Omicron subvariants emerged, with BA.2 becoming the most prevalent in May 2022, replacing the previously dominant BA.1 subvariant. As of September 2022, BA.5 is dominant in Japan^17^ and the data on the viral and clinical characteristics continue to be analyzed. Although vaccination against SARS-CoV-2 is in progress across the world, new variants and subvariants that evade SARS-CoV-2 neutralizing antibodies continue to be a threat.

As of April 2022, several therapeutic antiviral drugs against SARS-CoV-2 infection are available worldwide.^18-21^ In particular, remdesivir^18^ and molnupiravir,^20^ both ribonucleic acid (RNA) polymerase inhibitors, as well as nirmatrelvir, a SARS-CoV-2 3C-like (3CL) protease inhibitor, in combination with ritonavir as a pharmacokinetic booster,^19^ are antivirals recommended for treating patients at a high risk of developing severe illness. These agents have been approved for use in Japan as well.^22^ However, they were mostly approved prior to the emergence of the Omicron variant and were assessed in clinical trials in unvaccinated individuals.^18-21^ Therefore, little evidence exists for their efficacy against the Omicron variant and in vaccinated patients.^23^ In Japan, remdesivir has been approved for moderate-to-severe COVID-19, regardless of patient risk for progression to severe COVID-19, and for mild COVID-19 patients at a high risk of severe illness.^24^ However, treatment options for patients with mild-to-moderate COVID-19, regardless of vaccination status and risk of developing severe illness, are limited. Moreover, post-acute COVID-19 syndrome may affect anyone infected with SARS-CoV-2, even those with mild or no symptoms.^7, 8^ As a result, a new treatment or prevention measure for post-acute COVID-19 syndrome is needed.

Ensitrelvir fumaric acid (S-217622, hereafter ensitrelvir) is a novel oral SARS-CoV-2 3CL protease inhibitor that originated through collaborative research efforts between Shionogi & Co., Ltd., and Hokkaido University.^25^ Ensitrelvir has demonstrated antiviral efficacy against SARS-CoV-2, including the Omicron variant and other VOCs, in both *in vitro* and *in vivo* preclinical studies.^26-28^ The tolerability of 5-day administration of ensitrelvir has been confirmed in healthy adults in a phase 1 study (Japan Registry of Clinical Trials identifier: jRCT2031210202).^29^ Promising results have been observed in the ongoing phase 2/3 study comprising several components—phase 2a, phase 2b, phase 2b/3, and phase 3 (SCORPIO-SR) studies—designed to assess the antiviral efficacy of ensitrelvir. The phase 2a^30^ and 2b^31^ studies showed that once-daily ensitrelvir administered for 5 days demonstrated a significant reduction in viral titer and viral RNA compared with placebo in mild-to-moderate COVID-19. In the phase 2b study, while there was no significant difference between ensitrelvir and placebo in the time-weighted average change in total score corresponding to the 12 COVID-19 symptoms (with the exception of smell or taste disorder; **Supplemental Table 1**), planned and *post hoc* analyses demonstrated that ensitrelvir produced favorable improvements in a subtotal of 4 respiratory symptoms (planned) or in the composite of respiratory symptoms and feverishness (*post hoc*).^31^ Both studies demonstrated that ensitrelvir was safe and well tolerated.^30, 31^ The phase 2b study was conducted when Omicron was the dominant variant, and most patients had been vaccinated.^31^ These studies suggested the need to evaluate efficacy in endpoints that reflect clinical features of SARS-CoV-2 strains in circulation at the time.

In this study, we describe the protocol for a phase 3 study, with emphasis on the study rationale and key endpoints designed to assess the efficacy and safety of ensitrelvir in patients with mild-to-moderate COVID-19, irrespective of their risk status of developing severe illness and vaccination history.

## 2. Materials and methods

### 2.1. Study design

This is a multicenter, randomized, double-blind, placebo-controlled, phase 3 study to test the efficacy and safety of ensitrelvir in patients with mild-to-moderate COVID-19. The study will be conducted across several sites in Japan, Korea, Singapore, and Vietnam beginning in February 2022 and has been registered in the Japan Registry of Clinical Trials: jRCT2031210350. Patients will be randomized into 3 arms – ensitrelvir 125 mg, ensitrelvir 250 mg, and placebo. The intervention period will span 5 days (Days 1–5), during which patients will be administered ensitrelvir or placebo, followed by a follow-up period of 23 days (Days 6–28; **Figure 1**). Patients who provide consent to participate in the exploratory period (Days 29–337) will be further evaluated over the extended follow-up period.

**Figure 1.**
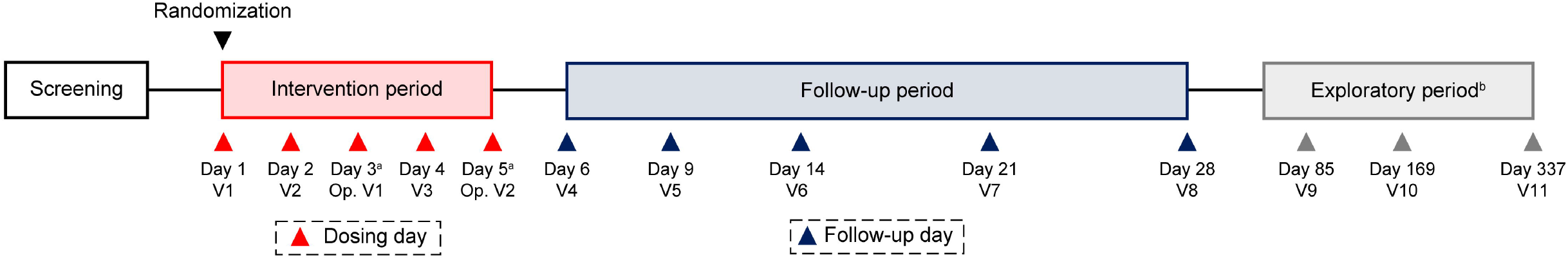
Study design. ^a^Optional visit. However, administration of study intervention and entry of patient diary should continue. ^b^Assessments will be performed only for patients providing consent/assent to participate in the exploratory period. Op = optional, V = visit.

### 2.2. Study ethics

This study will be conducted in accordance with the protocol and consensus ethical principles derived from the Declaration of Helsinki, the Council for International Organizations of Medical Sciences (CIOMS) international ethical guidelines, the International Council for Harmonisation of Technical Requirements for Pharmaceuticals for Human Use (ICH) Guidelines, and Good Clinical Practice guidelines. The protocol, protocol amendments, and other relevant documents will be submitted to the institutional review boards of the participating sites for approval. All patients will be required to provide written informed consent/assent to participate in the study before undergoing any evaluations or procedures. For minor patients, written informed consent will be obtained from a parent/legal guardian. All patients will be informed about the use of their personal, study-related data in accordance with data protection laws. The information and results of this study will be disclosed at the clinical study registration site and other sites. The results of this study may be published or presented at scientific meetings.

### 2.3. Randomization and blinding

Eligible patients will be randomly assigned to the 3 treatment arms in a 1:1:1 ratio using an interactive response technology, and a study intervention allocation table will be created. Randomization will be stratified by the time from the onset of COVID-19 to randomization (<72 hours vs. ≥72 hours) and by SARS-CoV-2 vaccination history (vaccinated [first dose completed] vs. unvaccinated). Placebo drugs will be indistinguishable in appearance, labeling, and packaging from treatment drugs. All relevant personnel, including the sponsor, patients, investigator, and sub-investigator, will remain blinded to the treatments. Unblinding at the request of the investigator may be permitted only in the event of an emergency or adverse event (AE) when knowledge of the intervention may be needed for an appropriate course of therapy.

### 2.4. Eligibility criteria

#### 2.4.1. Inclusion criteria

Patients must be capable of providing signed informed consent/assent and must be ≥12 years and <70 years of age at the time of providing informed consent/assent. All patients must (1) have been diagnosed as SARS-CoV-2–positive within 120 hours before randomization via either a nucleic acid detection test using a nasopharyngeal swab, nasal swab, or saliva (qualitative/quantitative reverse transcription-polymerase chain reaction [RT-PCR] test or an isothermal nucleic acid amplification method [e.g., the loop-mediated isothermal amplification method or the transcription-mediated amplification method]), a quantitative antigen test using a nasopharyngeal swab, nasal swab, or saliva, or a qualitative antigen test using a nasopharyngeal or nasal swab; (2) have a time from COVID-19 onset to randomization of ≤120 hours (when at least 1 of the 14 symptoms occurs: low energy or tiredness, muscle or body aches, headache, chills or shivering, feeling hot or feverish, stuffy or runny nose, sore throat, cough, shortness of breath, nausea, vomiting, diarrhea, smell disorder, and taste disorder); and (3) have at least 1 moderate symptom (COVID-19 symptom score 2; **Supplemental Table 1**) or severe symptom (COVID-19 symptom score 3; **Supplemental Table 1**) among the COVID-19 symptoms at enrollment (excluding smell or taste disorder and any symptoms present prior to COVID-19 onset) or at least 1 moderate symptom or severe pre-existing symptom (present prior to COVID-19 onset), which was considered to have worsened at baseline (before administration of study intervention). Patients’ body weight must be ≥40 kg (if patients are minors at the time of providing informed consent/assent). Additional inclusion criteria are described in **Supplemental Table 2**.

#### 2.4.2. Exclusion criteria

Patients will be excluded if they (1) have ≤93% (room air) saturation of percutaneous oxygen (SpO_2_) during wakefulness; (2) require oxygen administration or respirators; (3) are strongly suspected or expected to have worsening of symptoms associated with SARS-CoV-2 infection within 48 hours after randomization in the opinion of the investigator or sub-investigator; (4) have suspected active and systemic infections (excluding SARS-CoV-2 infection) requiring treatment at the time of randomization; (5) currently have or have a chronic history of moderate or severe liver disease or known hepatic or biliary abnormalities (with the exception of Gilbert’s syndrome or asymptomatic gallstones), or moderate or severe kidney disease (Grade 2 or higher based on Common Terminology Criteria for Adverse Events version 5.0^32^); (6) have used approved or unapproved drugs (e.g., interferon, convalescent plasma, monoclonal antibodies, immunoglobulins, antirheumatic drugs, corticosteroids [oral, injection, inhaled], ivermectin, or favipiravir) for the treatment of SARS-CoV-2 infection within 7 days prior to randomization; or (7) have used a strong cytochrome P450 3A (CYP3A) inhibitor, a strong CYP3A inducer, or products containing St. John’s wort within 14 days prior to randomization. Additional exclusion criteria are described in **Supplemental Table 2**.

Patients satisfying inclusion criteria 1–3 and not satisfying exclusion criteria 1–4 listed above are considered to have mild-to-moderate COVID-19.

### 2.5. Interventions

#### 2.5.1. Drug dose and administration

Based on the results of the phase 2a^30^ and 2b^31^ studies, patients will be orally administered a once-daily dosage of ensitrelvir 125 mg, ensitrelvir 250 mg, or a matching placebo. Two types of placebo doses will be administered: placebo-B, which is identical in appearance and packaging to ensitrelvir 125 mg, and placebo-D, which is identical in appearance and packaging to ensitrelvir 250 mg. For patients assigned to the placebo group, 3 tablets each of placebo-B and placebo-D will be administered on Day 1, followed by 1 tablet each of placebo-B and placebo-D administered on Days 2–5. For patients randomized to the ensitrelvir 125 mg group, a loading dose of 375 mg of ensitrelvir (3 tablets of 125 mg each) and 3 tablets of placebo-D will be administered orally on Day 1. Subsequently, a maintenance dose of ensitrelvir 125 mg (1 tablet of 125 mg) and 1 tablet of placebo-D will be administered orally once daily on Days 2–5. For patients randomized to the ensitrelvir 250 mg group, a loading dose of 750 mg of ensitrelvir (3 tablets of 250 mg each) and 3 tablets of placebo-B will be administered on Day 1. Subsequently, a maintenance dose of ensitrelvir 250 mg (1 tablet of 250 mg) and 1 tablet of placebo-B will be administered orally once daily on Days 2–5.

#### 2.5.2. Criteria for discontinuation

The study intervention will be discontinued for patients who experience worsening of SARS-CoV-2 infection, experience serious or intolerable AEs, are found to be ineligible for the study by the investigator or sub-investigator, request to discontinue the study interventions, are lost to follow-up, become pregnant, or for any other reason per the judgment of the investigator or sub-investigator.

#### 2.5.3. Prohibited concomitant therapy

Because ensitrelvir has an inhibitory effect on CYP3A, patients must refrain from consuming any foods and beverages containing grapefruit or Seville oranges and also avoid consuming products containing St. John’s wort during the study intervention period. Details of prohibited concomitant drugs are provided in the **Supplemental Appendix 1**.

### 2.6. Outcome measures and endpoints

The primary endpoint for this study will be the time to resolution of the 5 COVID-19 symptoms, defined as the time from the start of the study intervention until the resolution of the 5 symptoms (stuffy or runny nose, sore throat, cough, feeling hot or feverish, and low energy or tiredness) of SARS-CoV-2 infection. Patients will assess their symptoms using a partially modified index prescribed by the Food and Drug Administration (FDA)^33^ (**Supplemental Table 1**). Smell and/or taste disorder will be assessed using a 3-point scale ranging from 0 to 2, where 0 indicates the same as usual, 1 indicates less than usual, and 2 indicates no sense of smell or taste. The remaining 12 symptoms will be assessed using a 4-point scale ranging from 0 to 3, where 0 indicates no symptoms, 1 indicates mild symptoms, 2 indicates moderate symptoms, and 3 indicates severe symptoms. Resolution of symptoms will be assessed as follows: (1) for pre-existing symptoms that were present before the onset of COVID-19 and considered by the patient to have worsened at baseline, severe symptoms at baseline must have improved to moderate or better (including no symptoms), moderate symptoms at baseline must have improved to mild or better (including no symptoms), and mild symptoms at baseline must have remained mild or better (including no symptoms) and (2) for pre-existing symptoms that were present before the onset of COVID-19 and considered by the patient not to have worsened at baseline, severe symptoms at baseline must have remained severe or improved (including no symptoms), moderate symptoms at baseline must have remained moderate or improved (including no symptoms), and mild symptoms at baseline must have remained mild or better (including no symptoms). Symptoms other than the above (those not occurring before the onset of COVID-19 or those that occur at or after baseline) must have completely resolved. Patients will be considered to have achieved the primary endpoint if the 5 COVID-19 symptoms remain resolved for ≥24 hours.

The 2 key secondary endpoints for this study will be change from baseline on Day 4 in the amount of SARS-CoV-2 viral RNA (key secondary endpoint 1) and the time to first negative SARS-CoV-2 viral titer, defined as the time from the first administration of the study intervention until the first confirmation of SARS-CoV-2 viral titer below a predetermined detection limit (key secondary endpoint 2). To evaluate the antiviral and clinical efficacy more clearly, for the primary and key secondary endpoints, the primary analysis population was defined as patients randomized <72 hours from the onset of COVID-19.

Other secondary endpoints will include the time to resolution of the 5 COVID-19 symptoms without symptom recurrence, defined as no increase of any symptom scores to moderate or severe at least for 48 hours after achieving resolution; the time to resolution of the 12 and 14 COVID-19 symptoms; the proportion of patients with smell or taste disorder at each time point; the proportion of patients without resolution of COVID-19 symptoms 3 weeks after the study intervention; change from baseline in SARS-CoV-2 viral titer and viral RNA at each time point; and scores on the 8-point ordinal scale at each time point (**Supplemental Table 3**). Other outcomes will include time to first occurrence of smell and/or taste disorder defined as the time from the start of the study intervention to the first occurrence of smell and/or taste disorder. Additional endpoints are described in **Supplemental Table 2**. Safety assessments will include laboratory tests related to hematology, blood chemistry, coagulation, serology, urinalysis, and others (**Supplemental Table 4**); physical examination; and measurement of vital signs.

### 2.7. Rationale for the primary endpoint

The phase 2a^30^ and 2b^31^ studies demonstrated decreased viral load with ensitrelvir treatment. In the phase 2a and 2b studies, time to improvement of COVID-19 symptoms was used for clinical assessment, similar to the assessment of influenza treatment, and a significant difference was not observed between ensitrelvir and placebo arms. A more stringent endpoint such as the time to resolution of symptoms may be a meaningful clinical target for patients with SARS-CoV-2 infection, and the FDA also recommends the time to clinical recovery for COVID-19 treatment.^33^

In the phase 2a and 2b studies,^30, 31^ the 12 COVID-19 symptoms were assessed based on the FDA guidance.^33^ It was revealed after the initiation of the phase 3 study that stuffy or runny nose, sore throat, cough, feeling hot or feverish, and low energy or tiredness were the 5 most frequently observed symptoms in all 3 treatment groups of the phase 2b study conducted during the Omicron BA.1 epidemic (**Table 1**). Furthermore, these 5 COVID-19 symptoms were selected as the components of the primary endpoint in the phase 3 study after studying the clinical features of infection during the Omicron variant epidemic,^17, 34^ including the BA.1 and BA.2 epidemics, as they were expected to reflect the clinical characteristic of these epidemic variants and the subtypes.

**Table 1.**
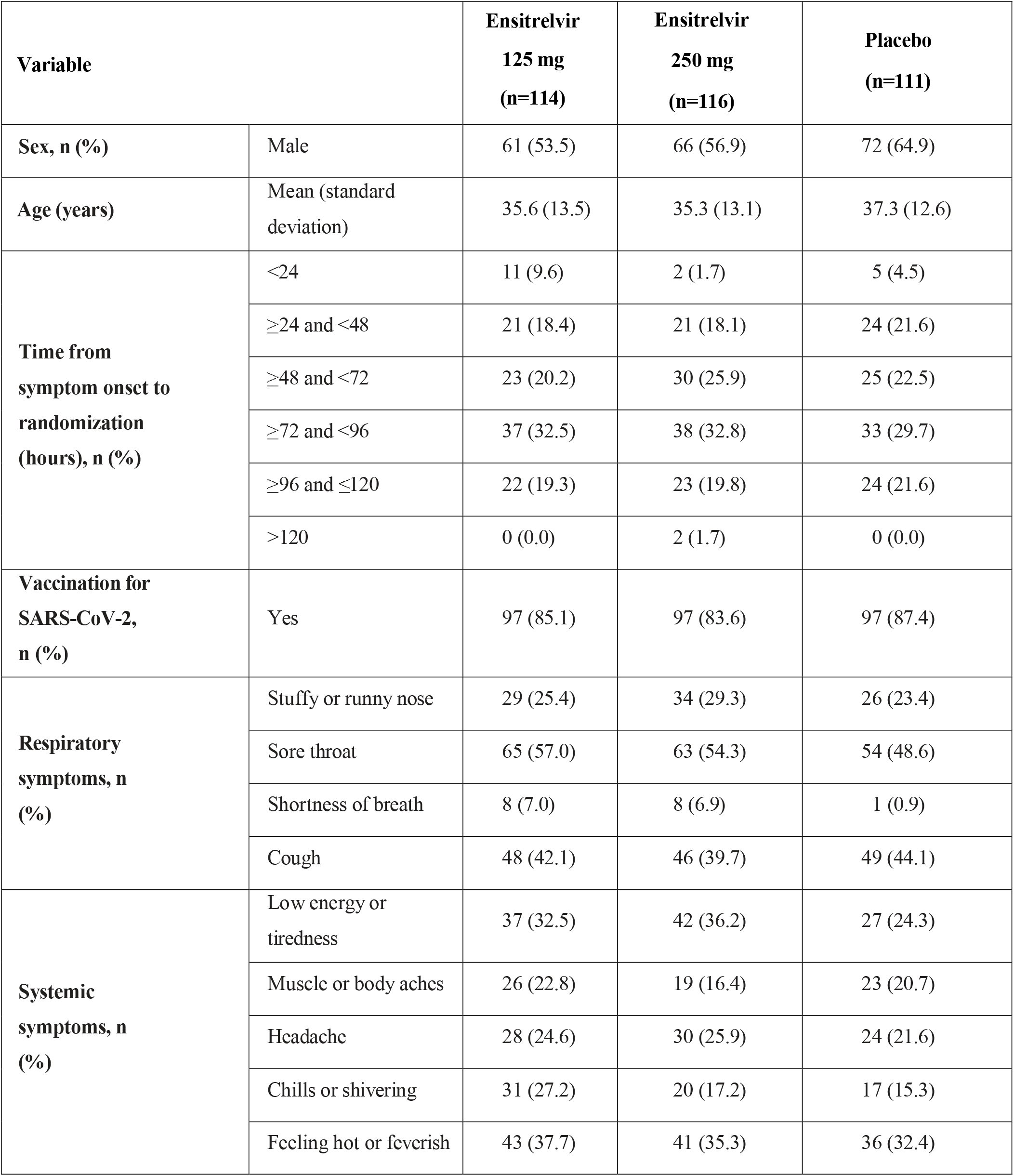
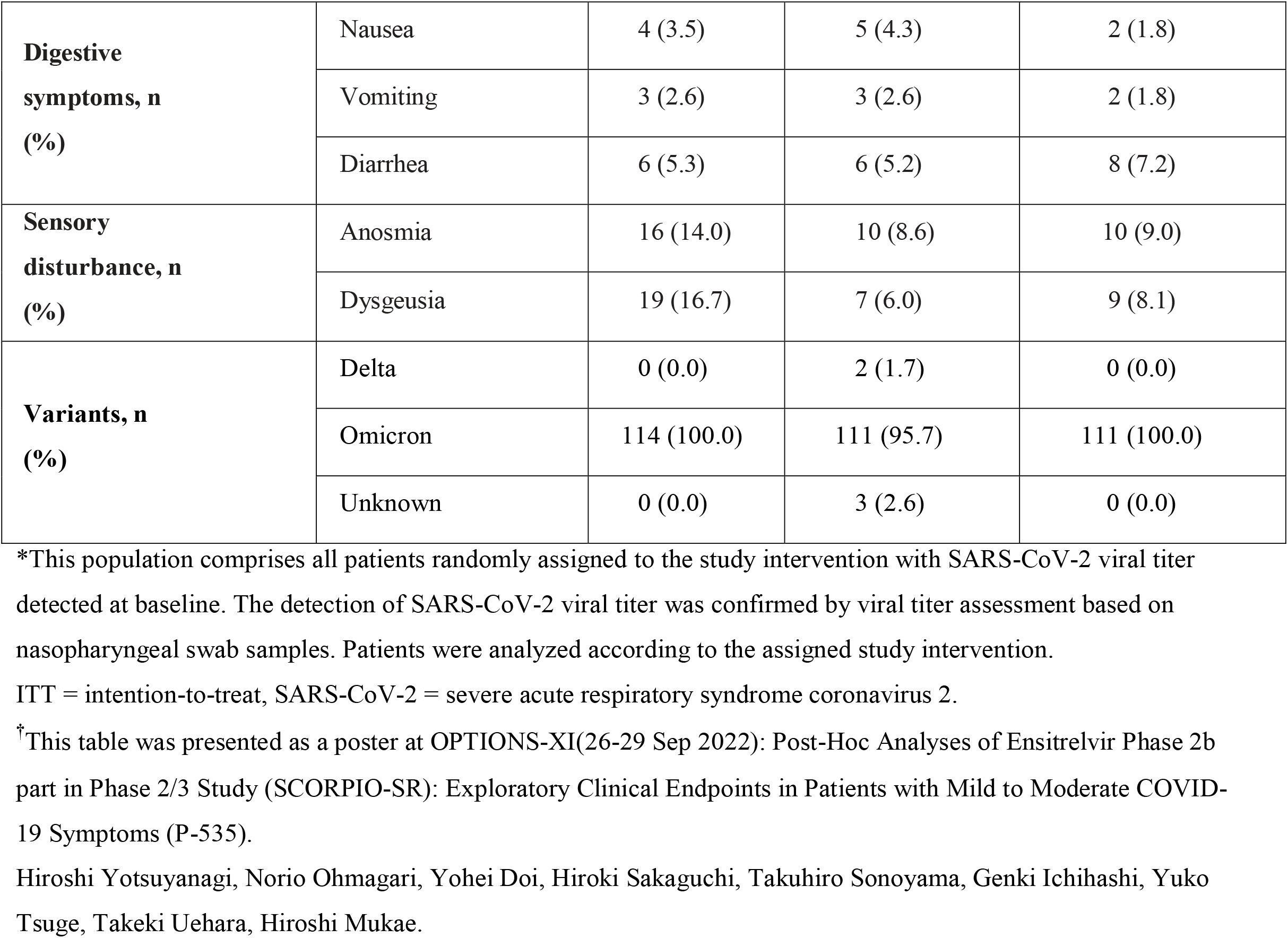
Patient demographics and clinical characteristics (ITT* population) in phase 2b study^†^.

The main pathophysiology of COVID-19 is due to viral proliferation for several days after onset followed by inflammatory reaction by the host immune system starting around 7 days after onset,^35^ and it is recommended to administer an antiviral drug in the earlier phase of infection. It has been reported that the patients with disease progression within 3 days and 4–7 days after onset of COVID-19 symptoms account for 31% and 37% of hospitalized COVID-19 patients, respectively, in the BA.1 and BA.2 epidemics in Japan, whereas the percentage has changed to 62% and 26%, respectively, in the BA.5 epidemic.^17, 34^ This observation suggests that the observation period to predict the prognosis has become shorter with newer subvariants and may support the importance of earlier antiviral treatment—<72 hours of onset of COVID-19 symptoms. Hence, we defined patients randomized <72 hours from the onset of COVID-19 as the primary analysis population to evaluate the antiviral and clinical efficacy more clearly. To confirm the overall ensitrelvir profile in this trial, we will also analyze the patients randomized within 120 hours from the onset of COVID-19.

In the *post hoc* analysis of the phase 2b study, the median time to resolution of the 5 COVID-19 symptoms in patients randomized to the study intervention <72 hours from the COVID-19 onset was shorter both in ensitrelvir 125 mg and 250 mg arms compared with the placebo arm (placebo, 10.4 days; ensitrelvir 125 mg, 6.9 days; ensitrelvir 250 mg, 6.3 days; **Figure 2**). Therefore, the time to resolution of the 5 symptoms of COVID-19 will be assessed as the primary endpoint to confirm the clinical benefit of antiviral treatment in patients with SARS-CoV-2 infection.

**Figure 2.**
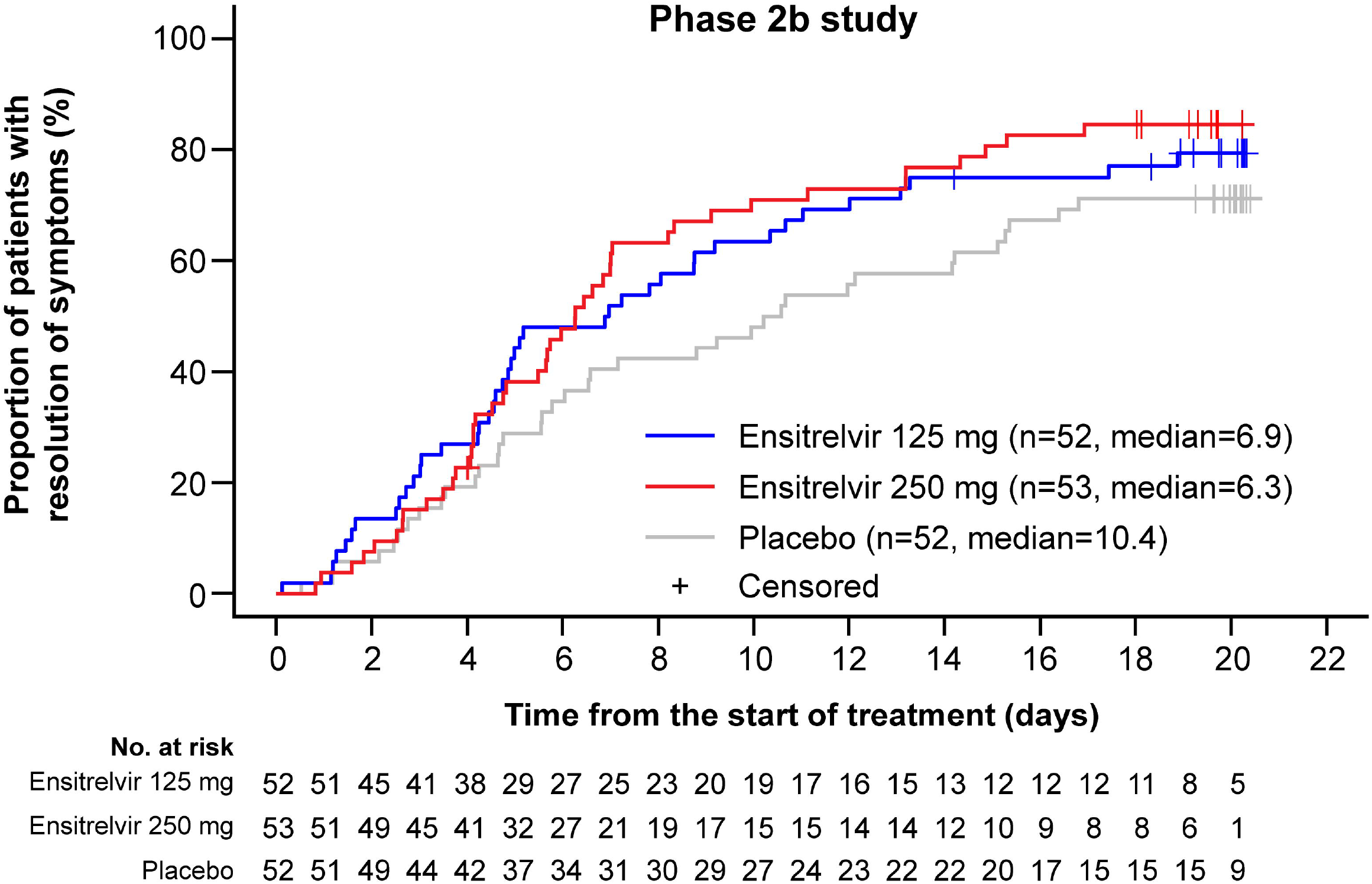
Kaplan-Meier plot of the time to resolution of the 5 COVID-19 symptoms in the patient group with <72 hours from the onset to randomization in the phase 2b study (ITT* population). *This population comprises all patients randomly assigned to the study intervention with SARS-CoV-2 viral titer detected at baseline. The detection of SARS-CoV-2 viral titer was confirmed by viral titer assessment based on nasopharyngeal swab samples. Patients were analyzed according to the assigned study intervention. COVID-19 = coronavirus disease 2019, ITT = intention-to-treat, SARS-CoV-2 = severe acute respiratory syndrome coronavirus 2.

The primary endpoint was finalized after consultations with experts and regulatory authorities.

### 2.8. Patient timelines

The schedule of activities for each patient will include efficacy and safety assessments (**Table 2**). Efficacy assessments will include virological examinations, patient diaries, and disease severity assessments. Patients will self-assess all 14 symptoms twice daily (morning and evening) from before the first dose of study intervention on Day 1 to Day 9 and once daily (evening) from Day 10 to Day 21. All endpoints assessed using the nasopharyngeal swab will be evaluated per the schedule of activities (**Table 2**). SpO_2_ levels and body temperature will be measured and recorded in the diary twice daily (morning and evening) from before the first dose of the study intervention on Day 1 until Day 9 and once daily (evening) from Day 10 to Day 21. If acetaminophen is taken as an antipyretic or analgesic, the COVID-19 symptom score will not be evaluated, and body temperature will not be measured until 4 hours after administration. Disease severity will be scored by the investigator or sub-investigator using the 8-point ordinal scale per the schedule of activities (**Table 2**). Safety assessments will be performed per the schedule of activities (**Table 2**).

**Table 2.**
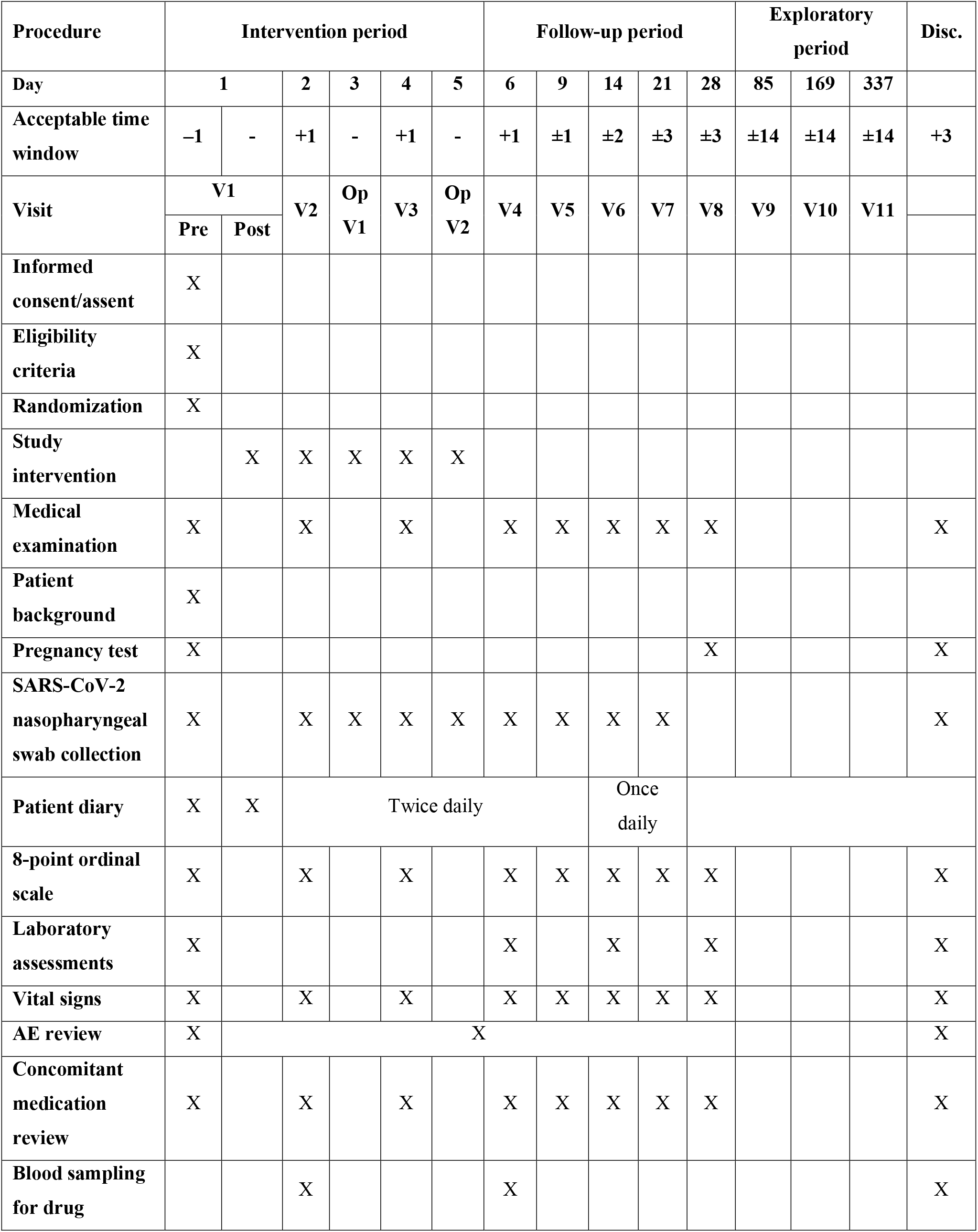
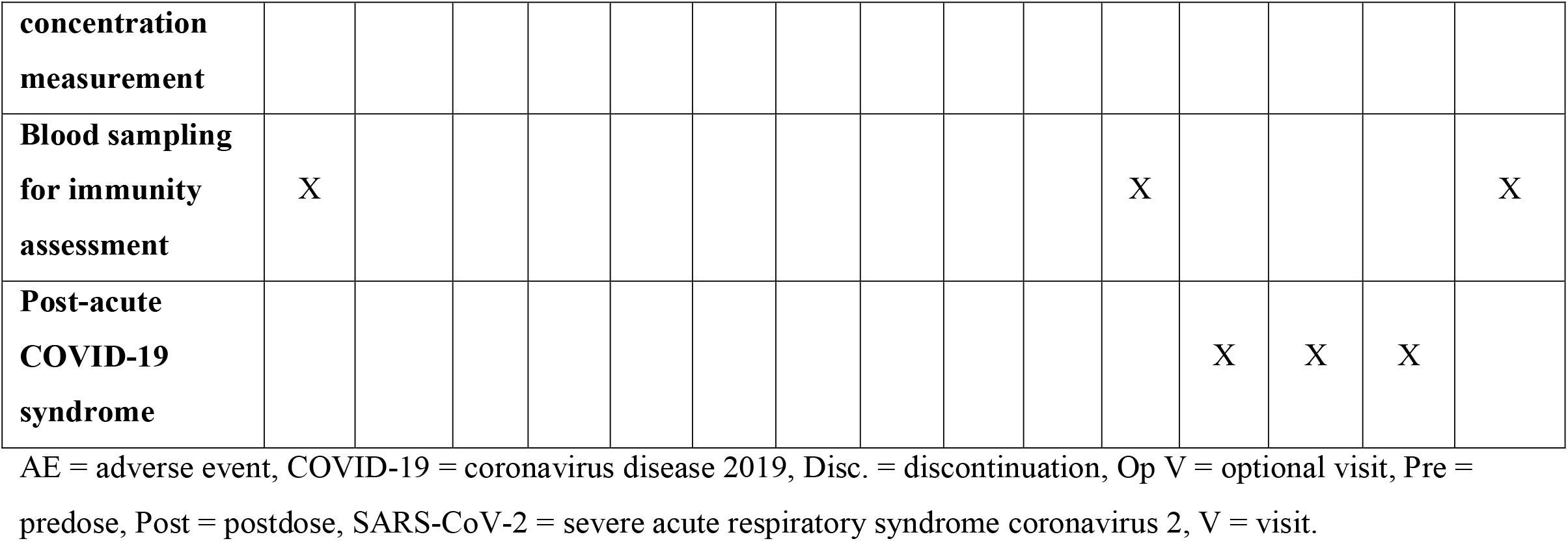
Schedule of activities.

### 2.9. Statistical analysis

#### 2.9.1. Analysis populations

Primary and secondary endpoints and other efficacy endpoints, except for endpoints related to the viral titer, will be assessed in the intention-to-treat (ITT) population, comprising all patients randomized to a study intervention and having SARS-CoV-2 infection confirmed by an RT-PCR test based on the nasopharyngeal swab sample from Visit 1 (pre-intervention). Secondary endpoints and other efficacy endpoints related to the SARS-CoV-2 viral titer will be assessed in the modified ITT (mITT) population, comprising all patients randomized to a study intervention with positive RT-PCR test results on Visit 1 and a SARS-CoV-2 viral titer detected at baseline. All safety assessments will be conducted in the safety analysis population (SAP) comprising patients who were randomized to a study intervention and who received at least 1 dose of the study intervention.

The population whose time from onset of COVID-19 to randomization in the ITT (or mITT for viral titer-based endpoints) population is <72 hours will be the primary analysis population for both the primary and key secondary endpoints. If a significant difference is found in the populations, the same analysis as the primary endpoint and key secondary endpoints will be performed on the entire ITT (or mITT) population.

Pharmacokinetic/pharmacodynamic analysis of the phase 2b study showed no clear difference in the antiviral efficacy between ensitrelvir 125 mg and 250 mg.^31^ As the 125 mg dose was suggested to achieve maximum antiviral effect, the target efficacy dose was set to 125 mg in the phase 3 study.

#### 2.9.2. Statistical tests

For discrete variables, summary statistics such as the number and proportion of patients in each group will be calculated. For continuous variables, summary statistics such as the number of patients, arithmetic mean (mean), standard deviation, minimum, median, and maximum values will be calculated for each group. The 95% confidence intervals (CIs) will be calculated using the Clopper-Pearson method^36^ and the bootstrap percentile method^37^ for the difference in median time. All statistical tests will be performed with a two-sided significance level of .05, unless otherwise specified. Note that the primary comparison will be conducted between the ensitrelvir 125 mg group and the placebo group, and comparison between the ensitrelvir 250 mg group and the placebo group will also be conducted in the same manner as a secondary comparison.

As primary analysis for the primary endpoint, a comparison of the time to resolution of the 5 COVID-19 symptoms will be performed between the ensitrelvir 125 mg group and placebo group using a Peto-Prentice generalized Wilcoxon test^38^ stratified by SARS-CoV-2 vaccination history at a one-sided significance level of .025 in the population with <72 hours from the onset of COVID-19 to randomization from the ITT population.

The primary analyses for each of the 2 key secondary endpoints are as follows: for the key secondary endpoint 1, a comparison of the change from baseline on Day 4 in the amount of SARS-CoV-2 viral RNA will be performed between the ensitrelvir 125 mg group and the placebo group using an analysis of covariance (ANCOVA^39^) with change from baseline in the amount of SARS-CoV-2 viral RNA as response and SARS-CoV-2 vaccination history and the amount of SARS-CoV-2 viral RNA at baseline as covariates at a one-sided significance level of .025 in the population with <72 hours from the onset of COVID-19 to randomization from the ITT population. For the key secondary endpoint 2, a comparison of the time to the first negative SARS-CoV-2 viral titer will be performed between the ensitrelvir 125 mg group and the placebo group using a log-rank test^38^ stratified by SARS-CoV-2 vaccination history at a one-sided significance level of .025 in the population with <72 hours from the onset of COVID-19 to randomization from the mITT population. In terms of type 1 error control induced by statistical tests for the primary analyses of primary endpoint and 2 key secondary endpoints, a fixed-sequence procedure will be applied in a hierarchical order of the primary endpoint, key secondary endpoint 1, and key secondary endpoint 2 in the comparisons between the ensitrelvir 125 mg group and the placebo group. In this procedure, if a preceding statistical test is not significant at a one-sided significance level of .025, the subsequent statistical tests will not be performed. If all statistical tests of the primary analyses for the ITT or mITT population with <72 hours from the onset of COVID-19 to randomization are significant, the same fixed-sequence procedure with the same hierarchical order will be applied to the key secondary analyses of the primary endpoint and the key secondary endpoints in the entire ITT or mITT population. In these key secondary analyses, time from COVID-19 onset to randomization (<72 hours, ≥72 hours) will be added as stratification factors or covariates.

Other analyses for the primary and key secondary endpoints in the population with <72 hours from COVID-19 onset to randomization from the ITT or mITT population are as follows: a comparison of the time to resolution of the 5 COVID-19 symptoms and the time to the first negative SARS-CoV-2 viral titer will be performed between the ensitrelvir 125 mg group and the placebo group using a log-rank test^38^ and Peto-Prentice’s generalized Wilcoxon test,^38^ respectively, stratified by SARS-CoV-2 vaccination history. Kaplan-Meier curves will be plotted for each treatment group. The median time to resolution of the 5 COVID-19 symptoms and first negative SARS-CoV-2 viral titer, along with the respective 95% CIs, will be calculated; the difference in median time between treatment groups, along with 95% CIs, will be calculated. The hazard ratio for the time to resolution of the 5 COVID-19 symptoms and the time to first negative SARS-CoV-2 viral titer of the ensitrelvir 125 mg group to the placebo group will be estimated using a Cox proportional hazards model^40^ stratified by SARS-CoV-2 vaccination history. Restricted mean survival time with a 21-day investigation period will be estimated for the time to resolution of the 5 COVID-19 symptoms and the time to first negative SARS-CoV-2 viral titer for each treatment group, and a comparison between the ensitrelvir 125 mg group and the placebo group will be performed. These analyses will be performed for the ITT or mITT population by adding time from COVID-19 onset to randomization (<72 hours, ≥72 hours) as stratification factors or covariates. Multiplicity adjustments will not be performed for the above analyses.

Analysis of the time to sustained resolution of the 5 COVID-19 symptoms for 48 hours or longer without any recurrence of the 5 COVID-19 symptoms for 2 days (48 hours) or longer after the resolution of the 5 COVID-19 symptoms will be performed using the same analysis as that for the primary endpoint (except for multiplicity adjustment).

For the analysis of the time to resolution of the 12 COVID-19 symptoms that persist for ≥24 hours between the ensitrelvir 125 mg and the placebo group, the same analyses as for the primary endpoint will be performed (except for multiplicity adjustment).

For analyzing the change from baseline in SARS-CoV-2 viral titer and the amount of viral RNA at each time point, least squares means and difference between the ensitrelvir 125 mg group and the placebo group based on the ANCOVA model^39^ will be calculated in the same manner as for the key secondary endpoint 1 (except for multiplicity adjustment). To evaluate the proportion of patients with a positive SARS-CoV-2 viral titer result at each time point and the proportion of patients with a score of ≥1, ≥2, ≥3, ≥4, ≥5, ≥6, and 7 on the 8-point ordinal scale at each time point, the Mantel-Haenszel test^41^ will be conducted to compare the ensitrelvir 125 mg group with the placebo group. The same stratification factors used for the primary endpoint will be applied. Additionally, for the proportion of patients with a score of ≥1, ≥2, ≥3, ≥4, ≥5, ≥6, and 7 on the 8-point ordinal scale at each time point, the risk ratio and risk difference between the ensitrelvir 125 mg group and the placebo group will be estimated using the Mantel-Haenszel method.^41^

#### 2.9.3. Sample size estimation

Referring to the Kaplan-Meier curve of the time to resolution of the 5 COVID-19 symptoms in a patient group with <72 hours from the onset of COVID-19 to randomization in the phase 2b study, we assumed a Weibull distribution as shown in **Figure 3**, considering the possibility of observing the effect conservatively in the ensitrelvir group. The assumed median time to resolution of the 5 COVID-19 symptoms corresponds to 8.3 days in the ensitrelvir 125 mg group and 11.1 days in the placebo group.

**Figure 3.**
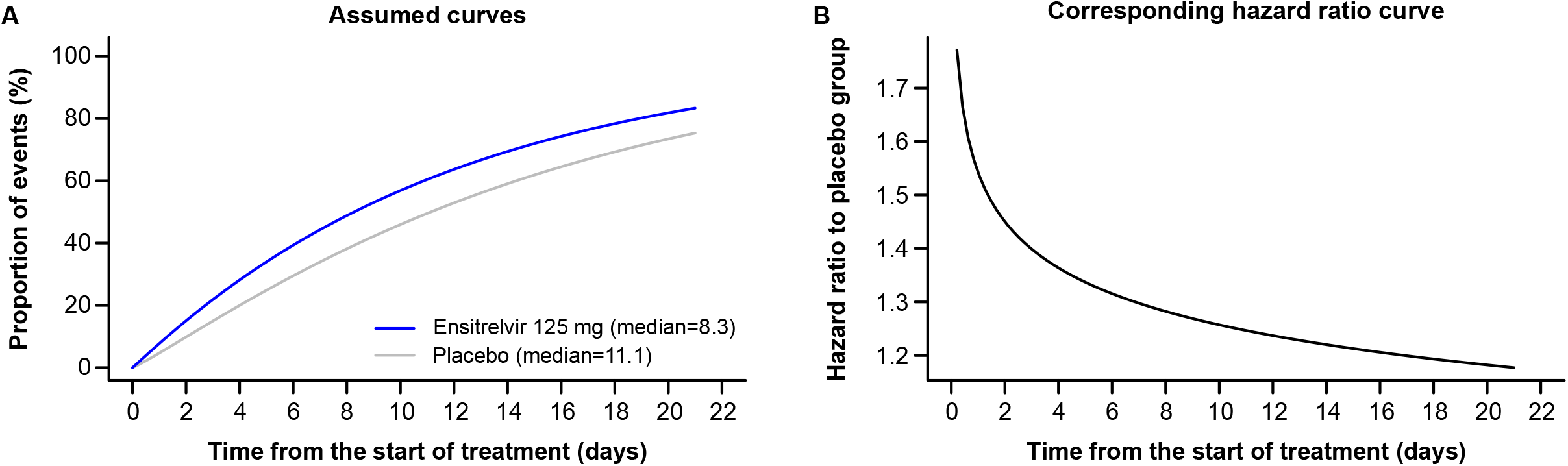
Weibull distribution (**A**) and corresponding hazard ratio curve (**B**) assumed when calculating the required number of patients for the phase 3 study for the time to resolution of the 5 symptoms of COVID-19. COVID-19 = coronavirus disease 2019.

The number of patients required to detect these differences with a power of 80% using the Peto-Prentice generalized Wilcoxon test^38^ with a one-sided significance level of .025 was calculated as 230 per group (the time from onset of COVID-19 to randomization being < 72 hours). Assuming about 10% of dropouts due to negative RT-PCR result in the test before enrollment, the number of patients to be enrolled with time from onset of COVID-19 to randomization of <72 hours was calculated as 260 per group for a total of 780 patients.

As described above, phase 3 consisted of 3 arms: the ensitrelvir 125 mg, ensitrelvir 250 mg, and placebo arms. The analysis population and the primary endpoint were changed based on the phase 2b outcomes as well as the sample size estimation. The original sample size estimation was as follows: in the phase 2b study, the median time to resolution for each ensitrelvir arm was conservatively assumed to be 8 days, while that for the placebo arm was assumed to be 10 days. The original primary comparisons were pairwise comparisons between each ensitrelvir group and the placebo group, which required multiplicity adjustment. Using the Bonferroni method^42^ for multiplicity adjustment, the sample size required to detect differences in median time to resolution with 80% power using the log-rank test^38^ at a one-sided significance level of .0125 was estimated to be 1428 patients (476 per group). Assuming a dropout rate of 10% due to a negative RT-PCR result before enrollment, the sample size of patients to be enrolled was estimated to be 1590 patients (530 per group).

#### 2.9.4. Imputation of missing data

Missing data will not be imputed; all statistical analyses will be based on observed cases, unless otherwise specified. Missing assessments in a patient’s diary after the initial administration of the study intervention will be imputed: morning times will be imputed as 11:59:59 and evening times will be imputed as 23:59:59 (only evening times apply for Days 10–21).

#### 2.9.5. Data management and monitoring

No interim analyses will be performed for this phase 3 study. An Independent Data Monitoring Committee will not be established. A Data and Safety Monitoring Board will be established for the purpose of third-party evaluation of safety throughout the study period. All patient data relating to the study will be recorded on electronic case report forms (eCRFs) unless electronically transferred to the sponsor or designee (e.g., laboratory data, electronic patient-reported outcomes). After the follow-up period is complete, all data in the eCRFs for each patient will be locked. Source data verification will be performed to confirm that the data entered into the eCRFs by authorized site personnel are accurate, complete, and verifiable from source documents, that the safety and rights of patients are being protected, and that the study is being conducted in accordance with the currently approved protocol and any other study agreements and applicable regulatory requirements.

All AEs and SAEs will be monitored from the time of providing informed consent/assent until the end of the follow-up period, as per the schedule of activities (**Table 2**). The investigator or sub-investigator or any qualified designee will be responsible for detecting, documenting, reporting AEs and SAEs, and will following up with patients for AEs and SAEs considered related to the study intervention or study procedures, or those that caused the participant to discontinue the intervention.

## 3. Discussion

We described the protocol for a multicenter, randomized, double-blind, placebo-controlled, phase 3 trial to assess the efficacy and safety of ensitrelvir, a novel oral SARS-CoV-2 3CL protease inhibitor, in expediting the resolution of symptoms in patients with mild-to-moderate COVID-19. We aimed to test the hypothesis that COVID-19 symptoms resolve sooner when patients are treated with ensitrelvir compared with placebo. Through this study, we sought to validate the potential antiviral efficacy of ensitrelvir observed in previous studies and to further establish its clinical efficacy and safety.

Approximately 81%–85% of patients with COVID-19 experience mild-to-moderate symptoms.^43, 44^ However, up to 14% of patients with mild-to-moderate illness can develop severe illness within a week.^43^ Specific groups of patients such as those above 75 years of age and those having comorbidities, such as obesity, chronic kidney disease, diabetes mellitus, hypertension, or heart failure, are predisposed to progressing from mild-to-moderate illness to critical forms.^43^ Therefore, several treatment strategies are directed toward protecting these vulnerable subgroups.^18-20^ Treating patients with mild disease early in the disease course is not only critical for preventing progression of the infection to severe forms but also for containing the spread of the virus.^43^ Treatments for individuals who are at a low risk of developing severe illness are limited, highlighting the need for the development of therapeutic interventions to treat such patients.

To maximize patient recruitment during the SARS-CoV-2 epidemic, the phase 3 study aims for a seamless study design with continuous patient enrollment after the phase 2b study.^31^ Therefore, the phase 3 protocol was amended based on data obtained in the phase 2b study^31^ and the clinical features of the epidemic caused by the Omicron variant and subvariants. However, all amendments, including the primary endpoint, analysis populations, statistical tests, and sample size estimation, were made blinded from the phase 3 data and were based on consultations with infectious disease experts and regulatory authorities.

Both the ensitrelvir 125 mg and 250 mg regimens demonstrated a potent antiviral efficacy against SARS-CoV-2 in the phase 2a^30^ and phase 2b^31^ studies, but a clear dose response was not observed between the 2 groups. Furthermore, analysis of the relationship between ensitrelvir exposure and viral titer response suggested that nearly maximum effect could be achieved by 125 mg treatment (unpublished data; manuscript in preparation). Either ensitrelvir 125 mg or 250 mg was originally planned to be selected during the phase 3 study based on the phase 2b study^31^, and the primary population was eventually determined to be patients in the ensitrelvir 125 mg group in the phase 3 study; however, both the 125 mg and 250 mg doses will be evaluated through the phase 3 study to assess the safety profile of ensitrelvir. Only the 125 mg dose is being evaluated in the ongoing SCORPIO-HR study (ClinicalTrials.gov: NCT05305547).

To evaluate the clinical efficacy of ensitrelvir against the SARS-CoV-2 infection, we will apply the primary endpoint focusing on the 5 major symptoms of the SARS-CoV-2 infection. In the *post hoc* analyses of the phase 2b study data, a trend toward a difference in the time to resolution of symptoms was observed between the ensitrelvir groups and placebo group; the time to resolution of symptoms was shorter in the ensitrelvir groups than in the placebo group. Additional analysis of the phase 2b study on smell and taste disorder revealed that compared with placebo, ensitrelvir reduced the number of patients experiencing both smell disorder (ensitrelvir 125 mg, *P* < .001; ensitrelvir 250 mg, *P* = .10; **Figure 4A**) and taste disorder (ensitrelvir 125 mg, *P* < .001; ensitrelvir 250 mg, *P* = .07; **Figure 4B**).

**Figure 4.**
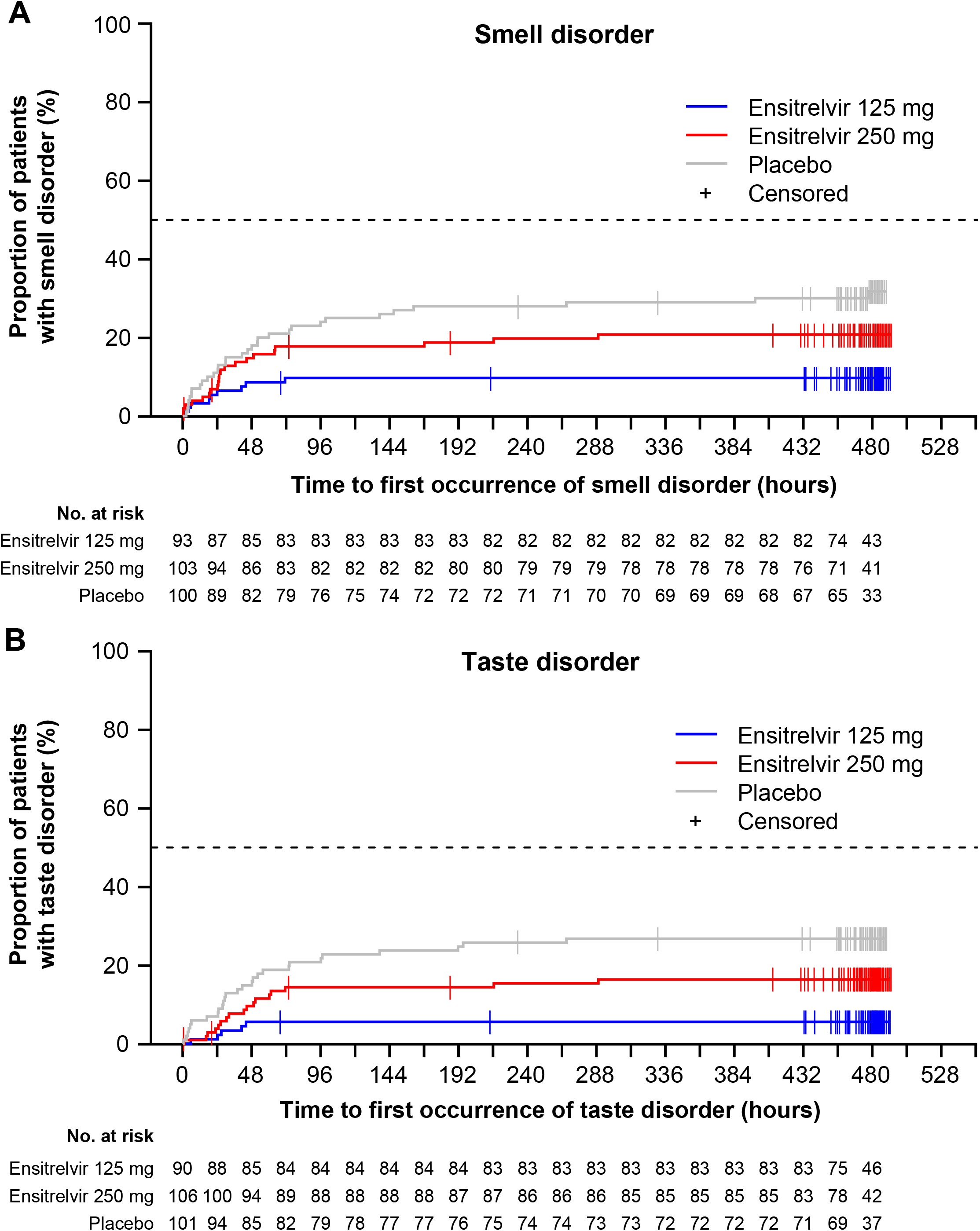
Proportion of patients showing occurrence of smell disorder (**A**) and taste disorder (**B**) in the phase 2b study (ITT* population). *This population comprises all patients randomly assigned to the study intervention with SARS-CoV-2 viral titer detected at baseline. The detection of SARS-CoV-2 viral titer will be confirmed by viral titer assessment based on nasopharyngeal swab sample. Patients will be analyzed according to the assigned study intervention. ITT = intention-to-treat, SARS-CoV-2 = severe acute respiratory syndrome coronavirus 2.

Irrespective of risk factors and the severity of infection, symptoms of COVID-19 may persist for a prolonged duration. Therefore, a stringent endpoint such as the time to resolution of symptoms may be suitable for evaluating the clinical efficacy of ensitrelvir in mild-to-moderate infections. Moreover, this endpoint is also appropriate in light of disease modification due to vaccination and the emergence of new variants. Since there is no information on how symptoms in mild-to-moderate COVID-19 progress over time, the endpoint of time to resolution of COVID-19 symptoms will provide clarity on how symptoms resolve with or without an antiviral treatment.

One of the strengths of this study is that it includes patients with COVID-19, irrespective of whether they are at a high risk of developing severe illness, which broadens its relevance to the general adolescent and adult populations. Additionally, this study will enroll both vaccinated and unvaccinated individuals, unlike other studies assessing oral antivirals that mainly recruited unvaccinated patients. Since vaccination has a large impact on how the infection manifests in individuals, including vaccinated patients in the study will provide a comprehensive understanding of the antiviral treatment that reflects the current status of the SARS-CoV-2 pandemic.

It is hoped that this phase 3 study will be able to confirm the clinical efficacy and safety of ensitrelvir in the treatment of mild-to-moderate COVID-19, and that the results will help provide evidence on symptom progression or improvement.

## Data Availability

All data produced in the present study are available upon reasonable request to the authors

## Abbreviations

3CL: 3C-like
AE: adverse event
ANCOVA: analysis of covariance
CI: confidence interval
CIOMS: Council for International Organizations of Medical Sciences
COVID-19: coronavirus disease 2019
CYP3A: cytochrome P450 3AFDA = Food and Drug Administration
eCRF: electronic case report form
ICH: International Council for Harmonization of Technical Requirements for Pharmaceuticals for Human Use
ITT: intention-to-treat
mITT: modified intention-to-treat
RNA: ribonucleic acid
RT-PCR: reverse transcription-polymerase chain reaction
SAE: serious adverse event
SAP: safety analysis population
SARS-CoV-2: severe acute respiratory syndrome coronavirus 2
SpO_2_: saturation of percutaneous oxygen
VOC: variant of concern
WHO: World Health Organization

## Acknowledgements

We thank Shintaro Tanaka, Masahiro Kinoshita, Satoshi Kojima, and Manami Yoshida for their contributions toward the development and review of this manuscript. Medical writing and editorial support were provided by Varsha Sreenivasan, PhD, of Cactus Life Sciences (part of Cactus Communications Pvt. Ltd.) and funded by Shionogi & Co., Ltd.

## Author contributions

Conceptualization: All authors.

Data curation: Hiroshi Yotsuyanagi, Norio Ohmagari, Yohei Doi, Genki Ichihashi, Takao Sanaki, and Hiroshi Mukae

Formal analysis: Takumi Imamura and Takao Sanaki

Visualization: Takumi Imamura

Methodology: Takao Sanaki

Supervision: Takumi Imamura, Takuhiro Sonoyama, Yuko Tsuge, Takeki Uehara, and Hiroshi Mukae

Project administration: Genki Ichihashi, Yuko Tsuge, and Takeki Uehara

Writing–-review and editing: All authors.

## Data availability

Data sharing is not applicable to this article as no datasets were generated or analyzed during the current study.

## SUPPLEMENTAL TABLE LEGENDS

**Supplemental Table 1**. COVID-19 symptom score.

^a^For items 1–10, sample item wording could be: “What was the severity of your symptom at its worst over the last 24 hours?”

^b^Score values are included in the table for ease of reference. The FDA cautions against including the score values within the response options presented to patients to avoid confusion.

COVID-19 = coronavirus disease 2019, FDA = Food and Drug Administration.

**Supplemental Table 2**. Additional eligibility criteria and endpoints.

AUC = area under the curve, EQ-5D-5L = EuroQol 5 dimension 5 level, RNA = ribonucleic acid, RT-PCR = reverse transcription-polymerase chain reaction, SARS-CoV-2 = severe acute respiratory syndrome coronavirus 2, SpO_2_ = saturation of percutaneous oxygen.

**Supplemental Table 3**. The 8-point ordinal scale for assessing symptom severity.

**Supplemental Table 4**. List of safety laboratory assessments.

^a^All events of ALT ≥3×ULN, and bilirubin ≥2×ULN (>35% direct bilirubin) or ALT ≥3×ULN and INR >1.5, if INR is measured (which may indicate severe liver injury), must be reported as an SAE.

ALP = alkaline phosphatase, ALT = alanine aminotransferase, APTT = activated partial thromboplastin time, AST = aspartate aminotransferase, BUN = blood urea nitrogen, CCL = C-C motif ligand, CK = creatine kinase, CRP = C-reactive protein, GGT = gamma-glutamyltransferase, HDL-C = high-density lipoprotein cholesterol, IFN = interferon, IgG = immunoglobulin G, IgM = immunoglobulin M, IL = interleukin, KL-6 = Krebs von den Lungen 6, LDH = lactate dehydrogenase, LDL-C = low-density lipoprotein cholesterol, MCH = mean corpuscular hemoglobin, MCV = mean corpuscular volume, PT/INR = prothrombin time and international normalized ratio, SAE = serious adverse event, TARC = thymus and activation-regulated chemokine, TG = triglycerides, UIBC = unsaturated iron binding capacity, ULN = upper limit of normal.

