## Supplemental Table 1 for "A Phase 2/3 study of S-217622 in participants with SARS-CoV-2 infection (Phase 3 part)"

**Supplemental Table 1.** COVID-19 symptom score.

| **S. No.** | **Symptom^a^** | **Response^b^** |
| --- | --- | --- |
| **1.** | Low energy or tiredness | 0: None  1: Mild  2: Moderate  3: Severe |
| **2.** | Muscle or body aches |  |
| **3.** | Headache |  |
| **4.** | Chills or shivering |  |
| **5.** | Feeling hot or feverish |  |
| **6.** | Stuffy or runny nose |  |
| **7.** | Sore throat |  |
| **8.** | Cough |  |
| **9.** | Shortness of breath (difficulty breathing) |  |
| **10.** | Nausea (feeling like you wanted to throw up) |  |
| **11.** | Vomiting (throw up) |  |
| **12.** | Diarrhea (loose or watery stools) |  |
| **13** | Rate your sense of smell in the last 24 hours | 0: My sense of smell is **the same as usual**  1: My sense of smell is **less than usual**  2: I have **no** sense of smell |
| **14.** | Rate your sense of taste in the last 24 hours | 0: My sense of taste is **the same as usual**  1: My sense of taste is **less than usual**  2: I have **no** sense of taste |

^a^For items 1–10, sample item wording could be: “What was the severity of your symptom at its worst over the last 24 hours?”

^b^Score values are included in the table for ease of reference. The FDA cautions against including the score values within the response options presented to patients to avoid confusion.

COVID-19 = coronavirus disease 2019, FDA = Food and Drug Administration.
