## Supplemental Table 2 for "A Phase 2/3 study of S-217622 in participants with SARS-CoV-2 infection (Phase 3 part)"

**Supplemental Table 2.** Additional eligibility criteria and endpoints.

| **Additional eligibility criteria** | |
| --- | --- |
| **Inclusion criteria** | **Exclusion criteria** |
| - Both male and female patients will be eligible. - Male patients must not donate sperm during the study intervention period and for at least 10 days after the administration of the last study intervention. - Male patients must practice abstinence or use contraception during the study period and for at least 10 days after the administration of the last study intervention. - Female patients must either not be of child-bearing potential or be pregnant or breastfeeding. - Women of child-bearing potential must use contraception/barriers during the study intervention period and for at least 10 days after the administration of the last study intervention, must not donate eggs during the study, and must have a negative result on a pregnancy test (urine or serum) within 24 hours prior to the first dose of the study intervention. | - Patients have previously received ensitrelvir. - Patients have donated ≥400 mL of blood in the 12 weeks or ≥200 mL blood in the 4 weeks prior to providing informed consent/assent. - Patients have been exposed to ≥4 new chemical entities within 12 months prior to dosing. - Patients are enrolled or have participated in any other clinical study involving an interventional drug or any other medical research within 28 days prior to providing informed consent. - Patients have difficulty in entering details into the patient diary properly due to cognitive decline, have a history of drug abuse, or are considered ineligible for the study by the investigator or sub-investigator due to sensitivity to any of the study interventions or their components thereof, have known allergic reactions to any drug, or have a history of other allergies (except for seasonal allergies), or for any other reason. |
| **Additional endpoints** | |
| - Time to resolution of fever (<37°C) - Time to sustained negative SARS-CoV-2 viral titer - Proportion of patients with positive SARS-CoV-2 viral titer at each time point - SARS-CoV-2 viral titer at each time point - Relative change rate from baseline in SARS-CoV-2 viral titer at each time point - AUC of change in SARS-CoV-2 viral titer - Proportion of patients with positive RT-PCR result at each time point - Amount of SARS-CoV-2 viral RNA at each time point - Relative change rate from baseline in the amount of SARS-CoV-2 viral RNA at each time point - AUC of change in the amount of SARS-CoV-2 viral RNA - Time to score of ≥1, ≥2, ≥3, ≥4, ≥5, ≥6, and 7 on the 8-point ordinal scale - SpO_2_ at each time point - Change from baseline in EQ-5D-5L - Ensitrelvir plasma concentration (Days 2, 6) | |

AUC = area under the curve, EQ-5D-5L = EuroQol 5 dimension 5 level, RNA = ribonucleic acid, RT-PCR = reverse transcription-polymerase chain reaction, SARS-CoV-2 = severe acute respiratory syndrome coronavirus 2, SpO_2_ = saturation of percutaneous oxygen.
