## Supplemental Table 3 for "A Phase 2/3 study of S-217622 in participants with SARS-CoV-2 infection (Phase 3 part)"

**Supplemental Table 3.** The 8-point ordinal scale for assessing symptom severity.

| **Descriptor** | **Score** |
| --- | --- |
| Asymptomatic | 0 |
| Symptomatic, no limitation of activities | 1 |
| Symptomatic, limitation of activities | 2 |
| Hospitalized, no oxygen therapy | 3 |
| Hospitalized, with oxygen therapy (<5 L/min) | 4 |
| Hospitalized, with oxygen therapy (≥5 L/min) | 5 |
| Hospitalized, with ventilation | 6 |
| Death | 7 |
