## Supplemental Appendix for "A Phase 2/3 study of S-217622 in participants with SARS-CoV-2 infection (Phase 3 part)"

**Supplemental Appendix 1. Prohibited concomitant drugs**

Drugs to be avoided from the time of providing informed consent/assent until the end of the follow-up period (Day 28) or at the time of discontinuation include approved and unapproved drugs (e.g., interferons, convalescent plasma, monoclonal antibodies, immunoglobulins, antirheumatic drugs; oral, injectable, or inhaled corticosteroids; ivermectin, and favipiravir) for the treatment of SARS-CoV-2 infection; CYP3A substrates; antiviral, antibacterial, or antifungal drugs (excluding those for external use); antipyretic analgesics other than acetaminophen; antitussives and expectorants; combination cold remedy; and investigational drugs as part of other clinical trials.

Drugs to be avoided from the time of providing informed consent/assent until 10 days after the last study intervention administration or at the time of study discontinuation include strong CYP3A inhibitors or inducers; P-glycoprotein (P-gp) inhibitors; breast cancer resistance protein (BCRP) inhibitors; and substrates for P-gp, BCRP, organic anion transporter polypeptide (OATP) 1B1, OATP 1B3, organic cation transporter 1 (OCT1), organic anion transporter 3 (OAT3), and multidrug and toxin extrusion 1 (MATE1) transporters.
